# Assessing unused residual Pfizer-BioNTech COVID-19 vaccine: a community observational study

**DOI:** 10.1101/2021.08.03.21261569

**Authors:** Stephen Hubbard, Rajlaxmi Bais

**Affiliations:** Bainbridge Prepares, Bainbridge Island, WA

**Keywords:** COVID-19, COVID-19 vaccine, Vaccine, Vaccine wastage, Pfizer-BioNTech, Pfizer Vaccine, Vaccine shortage, Vaccine cost

## Abstract

Worldwide morbidity and mortality associated with Covid-19 are severe and ongoing. The Pfizer-BioNTech vaccine is said to be up to 95% effective against severe disease or death. We were able to demonstrate that an additional 9.8% of COVID-19 vaccine doses could theoretically be given if the residual vaccine within the reconstituted Pfizer vials after six doses are extracted were used. This could be achieved by aseptically combining this excess vaccine from multiple vials to achieve full 0.3ml doses.

**Methods:** An observational study was conducted in April, 2021, at a mass vaccine site run by a community volunteer organization on Bainbridge Island, Washington. We measured the amount of Pfizer-BioNTech COVID-19 vaccine that was left in 172 vials after six doses had been withdrawn per Centers for Disease Control (CDC) protocol.

**Results:** A total of 30.68 ml of leftover vaccine was measured and discarded as medical waste. 1,036 doses were given from these vials. An extra 102 doses theoretically could have been given using the residual vaccine in the vials. This would have resulted in 9.8% additional doses of COVID-19 vaccine without requiring new vials.

**Conclusion:** The ability to combine solution from reconstituted Pfizer vaccine vials to minimize waste and obtain additional doses of vaccine could result in an increase in the number of individuals that could be vaccinated worldwide without additional cost. Further studies to validate our findings are warranted. Clinical trials to study the feasibility, safety and efficacy of protocols using this excess vaccine should be considered.

## Introduction

A worldwide total of over 178 million COVID-19 cases and 3.84 million deaths associated with COVID-19 has been recorded. ^1^ Due to the effectiveness and availability of vaccines from Pfizer-BioNTech, Moderna, Johnson & Johnson, and others, the number of cases and deaths are declining in some countries. Pfizer-BioNTech makes the first COVID-19 vaccine to receive emergency use authorization in the United States after clinical trials demonstrated that the vaccine was up to 95% effective against severe disease or death. ^2^

The risk of continued outbreaks, disease spread, disruption and death to the worldwide community will continue until effective vaccines are globally deployed, available, accessible, and administered. ^3,7^ For example, India has a population of 1.36 billion, and only 48.4 million individuals (3.5% of the population) have been fully vaccinated. ^1^ As recently as May 2021, India faced a catastrophic COVID-19 humanitarian crisis with more than 20.2 million reported cases and more than 380,000 deaths. Hospitals were lacking supplies and beds, while health care workers and the general public were exhausted and fearful of the limited availability of the COVID-19 vaccine. ^4^ Relief efforts are aimed at improving the vaccination supply and distribution of vaccine. A wide disparity between wealthy and poor countries exists in vaccination rate, due in part to lack of available vaccine. 7

This observational study took place through the help of *Bainbridge Prepares* (BP), a local organization comprised of community medical and non-medical volunteers. BP began administering Covid-19 vaccinations in January 2021 with the Moderna vaccine, which comes pre-prepared for usage. Shortly thereafter, the Pfizer-BioNTech vaccine became available for our use. Because of the increased complexity of vaccine reconstitution with the Pfizer product, the Bainbridge Island Medical Reserve Corps (BIMRC, a branch of Bainbridge Prepares) assisted in vaccine preparation. The BIMRC consists of volunteers with medical backgrounds and licenses (i.e., physicians, registered nurses, veterinarians, and other emergency medical responders). BP administered over 26,000 vaccinations through mass vaccine centers.

The BIMRC assisted in preparing Pfizer vaccine for injection by diluting and mixing the vaccine, then withdrawing and administering doses using single-dose, disposable syringes. We soon discovered that a significant amount of unused vaccine remained in the vials after the usual six doses were withdrawn. We were unable to find data quantifying this amount in the literature. Because of the COVID-19 pandemic and the ongoing need for more vaccine, our group looked at the quantity of measurable residual vaccine that had to be discarded as “waste” after the sixth vaccine dose was withdrawn.

“Wastage” of vaccines is normally defined as doses that do not get administered and must be discarded. This can happen when vials spend too much time outside a given temperature range; when vials are opened but not used within a certain number of hours; when the vaccines are not used by their expiration dates; and when the full volume of doses in the vial is not used.^5^ Wastage rates of this type with Covid-19 vaccines are estimated to be extremely low in the United States (less than 1%). It is assumed that there will be some wastage in vaccination programs; the World Health Organization estimates that 50% of vaccines are wasted around the world. ^6^

While wastage of whole doses of Pfizer-BioNTech Covid-19 vaccine is rare, and vaccination programs pride themselves on the use of every available dose, there is another potential source of wastage that has not been addressed: the residual vaccine left in the vials after the final full 0.3ml dose is drawn up (this may be after five or six doses). During our vaccine preparation process, it became apparent that vaccine was available but not being used in shots. In our study, this vaccine was the measurable residual after *six* doses had been withdrawn from the vial.

The Pfizer-BioNTech vaccine is unique among the vaccines available in the U.S. in that it must be diluted before use. Each vial contains 0.45 ml of vaccine after thawing. This is diluted with 1.8 ml of sterile saline, to make a total of 2.25 ml. of vaccine to be administered intramuscularly. ^8^ This vaccine is withdrawn using aseptic technique into single-use 1.0 ml syringes in 0.3 ml doses. Normally, 6 doses can be removed for use and rarely, 5 or 7. Less than 0.3 ml can be recovered from the vial after that final dose. Current FDA and Pfizer regulations state to “discard vial when there is not enough vaccine to obtain a complete dose. Do not combine residual vaccine from multiple vials to obtain a dose.” Therefore, the residual vaccine is discarded with the vial as medical waste.

In this study, we recorded how much additional vaccine could be withdrawn after the sixth full dose (0.3ml) of vaccine was obtained.

### Technique

Eight BIMRC vaccine handlers at a time participated during two different 4-hour shifts. Vaccine handlers were all licensed and experienced in handling medications and syringes. Vaccine handlers received specialized training in the proper technique for diluting and withdrawal of the Pfizer vaccine. The vaccine preparation room was separated for security, sterility, and lack of distraction. Quality control was provided by a board-certified nurse practitioner and the site physician lead, both of whom had experience in vaccination with the Pfizer product. Individually prepared syringes were inspected to ensure correct dose, secure needle/syringe system, lack of significant bubbles, and discoloration before being sent to the vaccination area for administration.

For this study, the vaccine handlers were instructed to withdraw the first 5 doses normally into single-use 1.0 ml syringes which were calibrated in 0.01 ml increments. (VanishPoint syringe; Retractable Technologies, Inc, Little Elm, TX). On the 6^th^ dose the handler withdrew as much vaccine as possible into the syringe, while maintaining sterility. The amount of vaccine over 0.3 ml was measured and recorded. This excess was returned to the vial and the syringe with its 0.3 ml dose was sent with the first 5 doses for quality control and use in vaccination in the clinic.

## Results

### Day One: April 3, 2021

Seven handlers participated

> *Total Vials*: 77
>
> From 77 vials, 13.6 ml of additional measurable residual vaccine was recorded after the sixth dose. This is an average of 0.17 ml per vial.

### Day Two: April 10, 2021

Eight handlers participated

> *Total Vials*: 95
>
> From 95 vials, 18.28 ml of residual vaccine was recorded after the sixth dose or 0.19 ml per vial.

**Day One + Day Two Total Vials: 172**

**Total administered doses across two days: 1036**

**Additional Theoretical Doses from residual vaccine: 102 or 9**.**8%**

Day One residual vaccine (13.6ml) + Day Two (18.28) = 31.88ml. However, four of these vials yielded an additional complete 0.3 ml dose, therefore only 30.68 ml was recorded as measurable residual vaccine. This residual vaccine could have been divided into an extra 102 doses. This would have been 9.8% more doses than were given.

## Discussion

We could have increased our vaccinations by 9.8% if we had used all the measurable residual vaccine which was subsequently discarded. Other local vaccine centers have confirmed having this leftover vaccine but due to the prohibition on combining vaccine from different vials to create additional doses, this residual vaccine had to be discarded unused.

The rationale given by Pfizer for not using this excess vaccine is the fear of contamination by mixing vaccine from different vials. Presumably, this could lead to increased risk of infection. However, infection from this vaccine is rare and the vaccine handlers were well-versed in sterile technique, so this risk seems small.

Another theoretical concern is that excessive handling of the vaccine might damage the lipid nanospheres used to contain mRNA. Vaccine handlers are taught to gently mix the vaccine and not shake it to avoid this complication. However, the extra manipulation of vaccine in syringes might damage the vaccine and lower its efficacy. Hopefully, Pfizer could answer this question. We believe that protocols for the safe mixing of unused vaccine could probably be created.

One limitation of this study is the small sample size. However, given the uniformity of this mass-produced vaccine, our findings seem likely to apply to the Pfizer vaccine generally. Further trials are warranted to test our results.

A second limitation is the fact that there still remained some unobtainable vaccine. Handlers were asked to record any excess vaccine that could be withdrawn after the sixth dose. However, there was an unknown amount of vaccine left in the vials that could not be removed with our syringes. Some of the vaccine seems to remain on the sides of the vial or in crevices around the stopper. Therefore, the amount of recoverable residual vaccine might be greater if better techniques were found to withdraw it.

The clinical and financial impact of getting an extra 9.8% from the vaccine vials could be substantial. Pfizer says it expects to make enough for 3 billion shots in 2021. Pfizer has said it will make $26 billion in vaccine sales in 2021, which would make it the best-selling medicine ever. ^9^ President Biden recently announced the purchase of 500 million doses of Pfizer vaccine, much of which will presumably be used in vaccinations in the developing world. Combining and using the residual vaccine in the vials containing those doses could theoretically be used to give an extra 49 million doses at no extra cost. This is enough to vaccinate 24.5 million people. We recommend that clinical trials be started to test the feasibility, safety, and efficacy of using this heretofore wasted resource: the unused vaccine left in the vial after the last dose has been withdrawn.

## Data Availability

All data used in the study are available for review.

---

**Figure 1:**
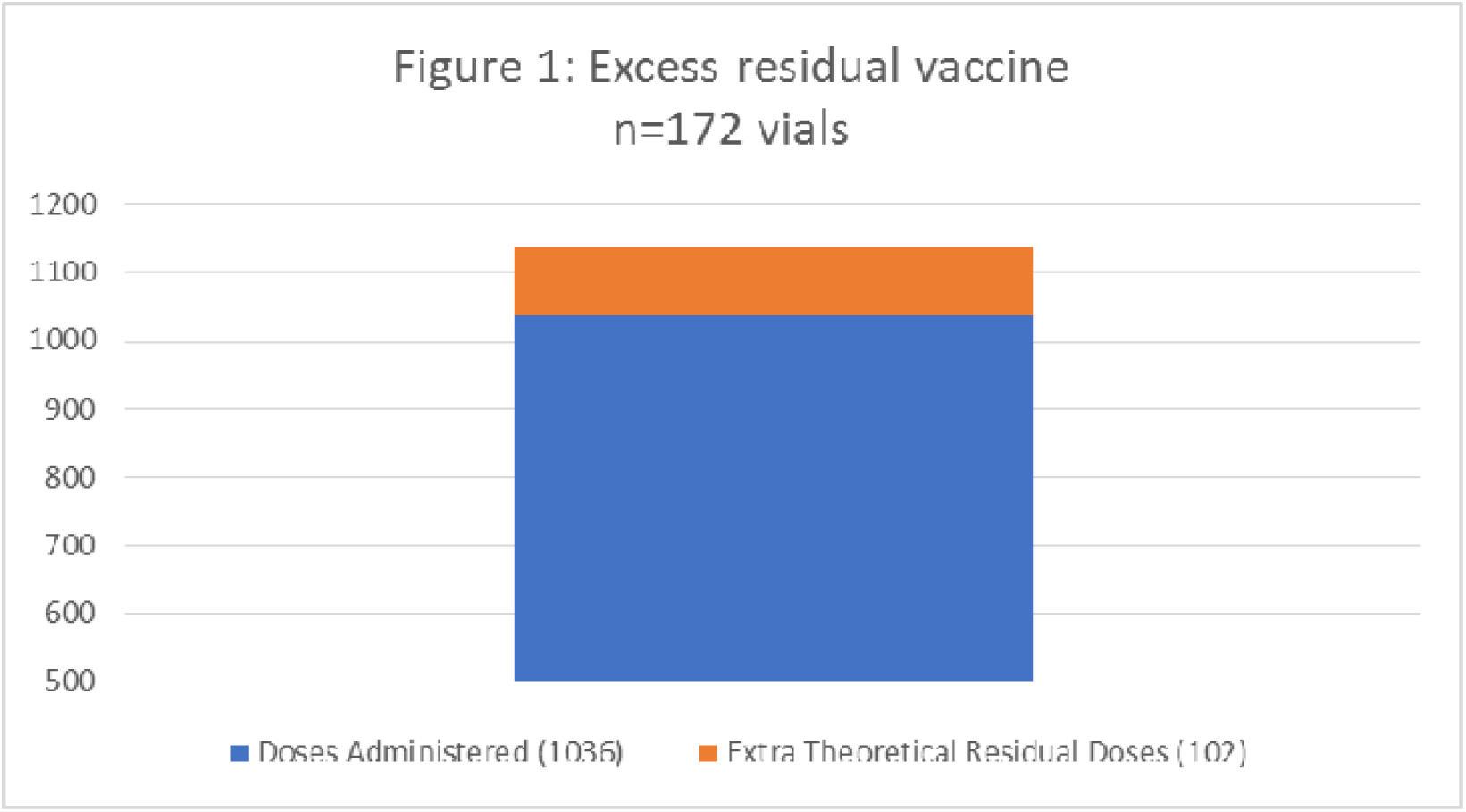
COVID-19 Vaccines Administered and Excess available residual COVID-19 Vaccine: Blue=Total number of vaccine doses administered (1036) Orange= Extra theoretical Residual doses that could have been given (102 doses or 9.8% additional, which would have been a total of 1138 doses)

## References

1. The New York Times Company (2021). New cases and deaths. https://www.google.com/search?q=total+death+toll+from+covid&rlz=1C1GCEV_enUS865US865&oq=total+death+toll+from+covid&aqs=chrome..69i57j0i457j0l8.2816j0j1&sourceid=chrome&ie=UTF-8

2. Katella, Kathy (2021). Comparing the COVID-10 Vaccines: How are they different? Yale Medicine. Retrieved from https://www.yalemedicine.org/news/covid-19-vaccine-comparison

3. Wouters, O.J., Shadlen, K.C., Salcher-Konrad, M., et al. (2021). Challenges in ensuring global access to COVID-19 vaccines: production, affordability, allocation, and deployment. The Lancet. 397 (10278),1023–1034. https://www.thelancet.com/journals/lancet/article/PIIS0140-6736(21)00306-8/fulltext

4. The Lancet (2021). India’s COVID-19 emergency. The Lancet. 397 (10286), 1683. https://www.thelancet.com/journals/lancet/article/PIIS0140-6736(21)01052-7/fulltext

5. Timsit, A. (2021). How many Covid vaccines go to waste? Quartz. https://qz.com/2013918/some-countries-are-wasting-more-covid-19-vaccines-than-others/

6. Schiffling, S. and Breen, L., (2021). COVID vaccine: some waste is normal-but here’s how it is being kept to a minimum. The Conversation. https://theconversation.com/covid-vaccine-some-waste-is-normal-but-heres-how-it-is-being-kept-to-a-minimum-152772

7. The New York Times Company (2021) https://www.nytimes.com/interactive/2021/world/covid-vaccinations-tracker.html

8. FDA (2021). Fact sheet for healthcare providers administering vaccine. https://www.fda.gov/media/144413/download#:∼:text=%E2%80%A2-,The%20Pfizer%2DBioNTech%20COVID%2D19%20Vaccine%20Multiple%20Dose%20Vial%20contains,and%20diluted%20prior%20to%20administration.

9. Rowland, C. (2021). Inside Pfizer’s race to produce the world’s biggest supply of covid vaccine. Washington Post. https://www.washingtonpost.com/business/2021/06/16/pfizer-vaccine-engineers-supply/

